# SARS-CoV-2 Breakthrough Infections among the Healthcare Workers Post-Vaccination with ChAdOx1 nCoV-19 Vaccine in the South Indian State of Kerala

**DOI:** 10.1101/2021.08.07.21261587

**Authors:** Aleena Issac, Jose J Kochuparambil, Linu Elizabeth

**Affiliations:** Clinical Quality Auditor, Department of Service Excellence and Quality, Mary Queen’s Mission Hospital, Kerala, India; Chief Quality Officer, Department of Service Excellence and Quality, Mary Queen’s Mission Hospital, Kerala, India; Infection Control Nurse, Department of Service Excellence and Quality, Mary Queen’s Mission Hospital, Kerala, India

**Keywords:** COVID-19, Vaccine effectiveness, Covishield vaccine, ChAdOx1 nCoV-19 Vaccine, Third wave

## Abstract

**Objective:** The national COVID-19 vaccination campaigns in India, prioritized healthcare workers to be vaccinated with ChAdOx1 nCoV-19 (AZD1222). So, we aimed to ascertain the effectiveness of ChAdOx1 nCoV-19 vaccine among the healthcare workers by characterizing the breakthrough infections which occurred 14 days after the second dose of the vaccine.

**Methods:** We prospectively evaluated the ‘Institutional Covid Vaccination Database’ to categorize the SARS-CoV-2 positive healthcare workers into 2 cohorts: infected vaccinated and infected unvaccinated cohort. Along with comparing the descriptive statistics of the cohorts, the relative risk (RR), relative risk reduction (RRR), and the vaccine effectiveness were calculated. The study population was compared for the significant difference by using the parametric model (binary logistic regression) for the Odds ratio and the non-parametric model (cox proportionality model) for the Hazard ratio.

**Results:** Out of the 324 healthcare workers employed in our institution 243 (75%) were vaccinated. When 6.58% (16) of the vaccinated healthcare workers were tested positive, 43.75% (35) were infected in the unvaccinated cohort and the median time of infection in the vaccinated cohort was 65 days (range 20 -91 days). While the cox proportionality model reveals that the age, sex, and the contact exposure status of the cohort are not significant factors for getting infected after being vaccinated, binary logistic regression proves that a significant relationship exists between the incidence of infection and the unvaccinated status (p = 0.001, OR = 4.278). Completing the two-dose of vaccination decreased the hazard of testing positive for the SARS-CoV-2 by 84.96 % when compared with the unvaccinated individuals, which is the effectiveness of the ChAdOx1 nCoV-19 vaccine.

**Conclusion:** There is an increased risk of the third wave of COVID-19 infection in the upcoming months. So, primary infection control measures should be followed even if vaccinated, to prevent the risk of breakthrough infections.

## Introduction

In India, the COVID-19 vaccination campaigns were originally conducted using the ChAdOx1 nCoV-19 vaccine (popularly known in India as Covishield) and BBB-152 (Covaxin) [1]. Healthcare workers and the frontline workers were prioritized in the initial phase of the vaccination campaign [2]. Earlier in May 2021, the vaccination opened to all individuals aged more than 18 years [3] and as of July 31, 2021, more than 10.35 crore Indians were fully vaccinated [4]

ChAdOx1 nCoV-19 (AZD1222) vaccine had been extensively studied and the overall efficacy more than 14 days after the second dose was found to be 66.7% (95% CI 57.4–74.0) and 70·4% (95·8% CI 54·8-80·6) respectively in two studies [5-6]. The vaccine effectiveness against hospitalization due to COVID-19 from day 22, after the first dose was 100%. This has provided an impetus to be authorized for use in several countries including India. Current data regarding the efficacy and the immunogenicity of the vaccine against the SARS-CoV-2 and the emerging variants propounds an overall efficacy of more than 65% in the majority of the studies [5-7].

The reduced risk of disease or infection among individuals which is attributed towards the vaccination in real-world conditions is termed as the effectiveness of the vaccine. World health organization has published standard interim guidelines for evaluating the effectiveness of COVID-19 vaccines [8]. According to the Ministry of Health and Family Welfare and ICMR, the breakthrough infection rates with the ChAdOx1 nCoV-19 vaccine were limited to 0.02% -0.04% [9]. Post-marketing real-life studies should be conducted rigorously to ascertain this level of effectiveness.

Healthcare workers had sustained exposure to the SARS-CoV-2 virus [10] and understanding the effectiveness of the vaccination in this cohort can be pivotal. So, we aimed to describe the COVID-19 vaccination status of the employees and to appraise the effectiveness of ChAdOx1 nCoV-19 vaccine by characterizing the breakthrough infections reported in a secondary care hospital (Mary Queen’s Mission Hospital, Kanjirappally) in the South Indian State of Kerala.

**SUMMARY BOX**

**What is already known on this subject?**

The vaccine being the final resort to fight against the COVID-19 pandemic, the incidence of breakthrough infections in fully vaccinated healthcare workers is quite distressing in the South Indian state of Kerala. World Health Organization states AZD1222 vaccine has 63.09% efficiency for symptomatic SARS-COV infection.

**What does this study add?**

The study is the foremost in evaluating vaccine effectiveness of 84.96% in fully vaccinated Healthcare workers in a Private Hospital of Kerala. The incidence of infections is more in the unvaccinated cohort (43.75%) than in the vaccinated cohort (6.58%). Factors like age, sex, and exposure status were found to have no relation to the incidence of breakthrough infections. Our study highlights the need for strict compliance of infection control practices even after vaccinations to prevent breakthrough infections.

## Methodology

The vaccination commenced on January 27, 2021, and the second dose was administered four to six weeks later from March 3, 2021. The Department of Service Excellence & Quality, Infection Prevention Wing of the hospital maintains a prospective COVID-19 database which includes the COVID-19 infection data, regarding the Reverse Transcriptase Polymerase Chain Reaction (RT-PCR) assay of the nasal swab and vaccination status of all the employees. All the employees were under surveillance after the vaccination process to quantify the breakthrough infections.

We conducted a single-center, prospective study by divided the groups into two cohorts [8]: infected vaccinated cohort and infected unvaccinated cohort (time frame of the study is presented in Figure 1). Breakthrough infections were defined as the COVID-19 infections occurring after 14 days of completing the primary series of vaccination. The symptomatic suspected cases, as well as the high-risk contacts of infected cases, were confirmed using RT-PCR screening with nasopharyngeal swabs. The data collection was conducted till July 15, 2021.

**Figure 1:**
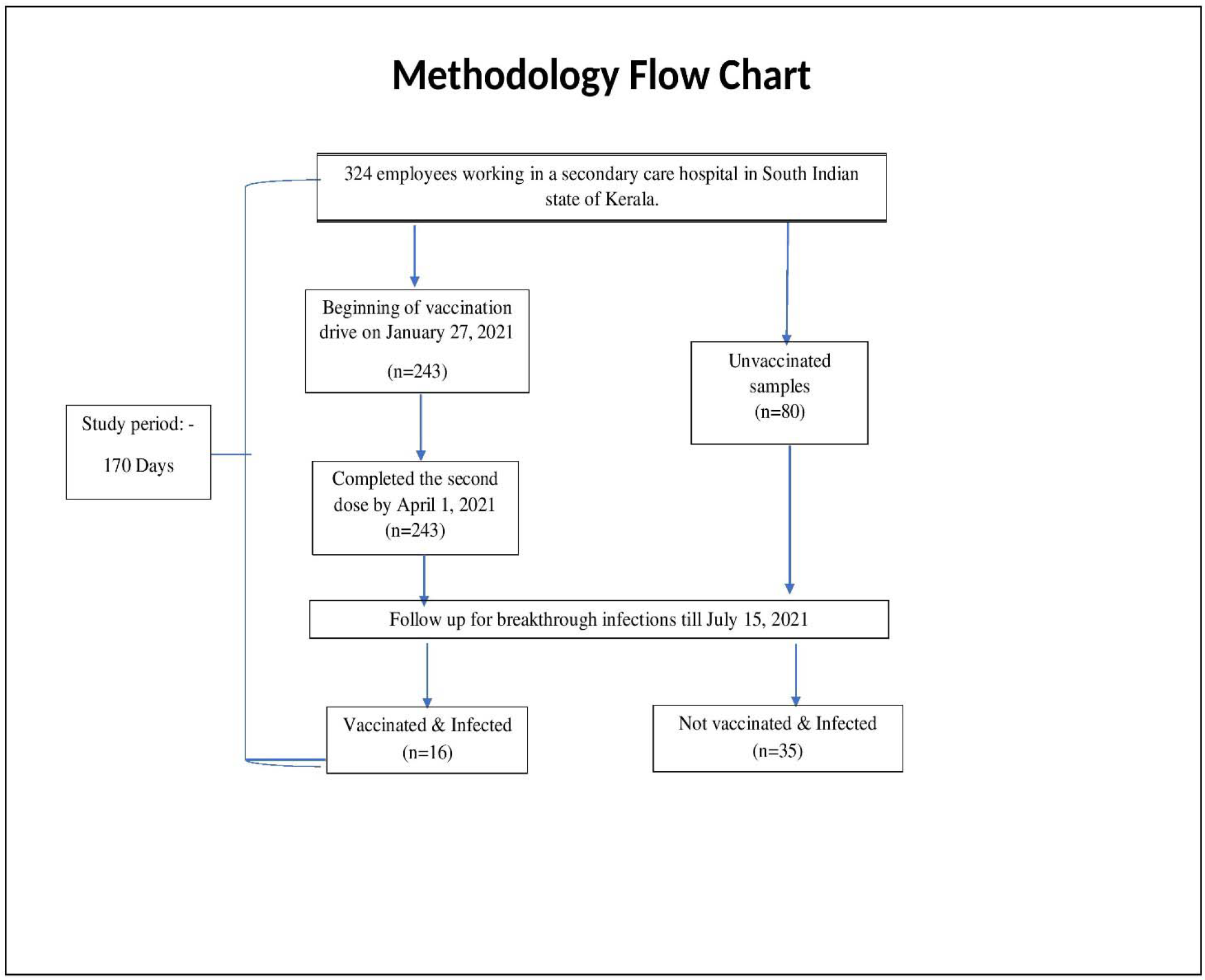
Selection and allocation of study sample into two cohorts – Vaccinated infected versus Not vaccinated infected

Data were analyzed using SPSS version 20. All the hypothesis tests were two-sided and evaluated at a significance level of 0.05. The descriptive statistics of age, sex, department, and contact exposure grade were compared between the cohorts. The symptomatic profile and prognosis of vaccinated positive healthcare workers were evaluated along with reasons for not being vaccinated. The relative risk (RR) and relative risk reduction (RRR), were calculated to evaluate the decreasing incidence of testing positive against the SARS-CoV-2 after vaccination. The vaccine effectiveness was calculated using the following formula [11]: -

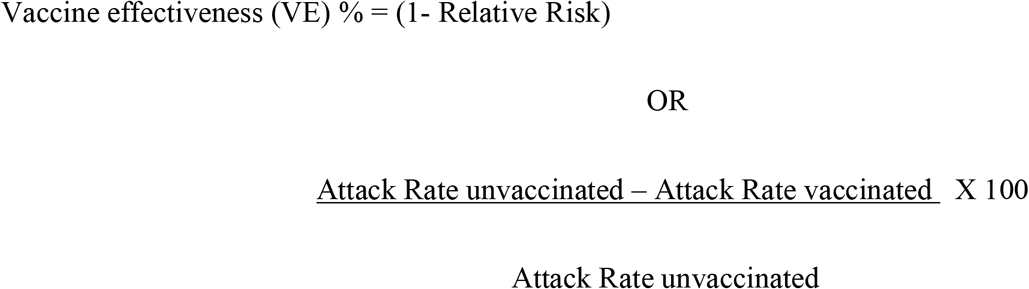

The cohorts are compared and checked for the significant difference by using the parametric model (binary logistic regression) and the non-parametric model (time-dependent cox proportionality hazard model). The event time observed was the number of days from the second dose until a COVID-19 infection was confirmed and the event was COVID-19 infection. Covariates tested were age, sex, and contact exposure status.

### Research Ethics Approval

Ethical approval is granted by the Institutional Ethical Committee of Mary Queen’s Mission Hospital (MQMH/IRB/21-23-00) upon ensuring the confidentiality of the personal information of the Hospital staff. Written consent was waived and verbal consent is obtained prior to the data collection.

## Results

We prospectively evaluated the 324 employees working in our hospital throughout the study period of 170 days (January 27, 2021 -July 15, 2021). Of this population, 243 (75%) completed the full primary cycle of vaccination, i.e., completed 14 days post-vaccination with 2 doses of ChAdOx1 nCoV-19 vaccine, and 80 (25%) were not vaccinated. Reasons for not being vaccinated (80) include breastfeeding 33.75% (27), pregnancy 13.75% (11), expecting pregnancy 15% (12), previous history of allergic reactions 0.0375% (3), and vaccine hesitancy 33.75% (27).

All the fully vaccinated staff received their second dose within 50 days of receiving the first dose of the vaccine. The demographic details of the vaccinated and the unvaccinated groups along with the details of these groups pooled together are described in table 1.

**Table 1:** Demographic details of the total study population as well as the vaccinated and unvaccinated cohort.

| **Demographic Characteristics** | **n=324** | **Vaccinated (n=243)** | **Unvaccinated (n=80)** |
| --- | --- | --- | --- |
| **Mean Age (SD)** | 34.09 (± 9.43) | 35.28 (± 10.02) | 30.26 (± 6.26) |
| **Sex** |  |  |  |
| Female | 272 (83.95%) | 194 (79.83%) | 77 (96.25%) |
| Male | 52 (16.04%) | 49 (20.16%) | 3 (3.75%) |
| **Department/Unit** |  |  |  |
| Doctors | 18 (5.55%) | 18 (7.4%) | 0 |
| Nurses | 165 (50.92%) | 106 (43.62%) | 59 (73.75%) |
| Pharmacist | 17 (5.24%) | 16 (6.58%) | 1 (1.25%) |
| Physiotherapist | 4 (1.23%) | 4 (1.64%) | 0 |
| Dietician | 1 (0.3%) | 1 (0.41%) | 0 |
| Administration | 8 (2.46%) | 8 (3.29%) | 0 |
| Accounts & Finance | 24 (7.4%) | 22 (9.05%) | 2 (2.5%) |
| Medical Records Department | 4 (1.23%) | 3 (1.23%) | 1 (1.25%) |
| Lab & Diagnostics | 50 (15.43%) | 39 (16.04%) | 11 (13.75%) |
| Housekeeping | 23 (7.09%) | 18 (7.4%) | 5 (6.25%) |
| Biomedical Engineering | 1 (0.3%) | 0 | 1 (1.25%) |
| Quality Department | 4 (1.23%) | 4 (1.64%) | 0 |
| Maintenance | 4 (1.23%) | 4 (1.64%) | 0 |
| Ambulance Driver | 1 (0.3%) | 1 (0.41%) | 0 |
| **Infection Status** |  |  |  |
| Infected | 51 (15.74%) | 16 (6.58%) | 35 (43.75%) |
| Not Infected | 273 (84.25%) | 227 (93.41%) | 46 (57.5%) |

A total of 51 employees tested positive for COVID-19 by RT-PCR from the beginning of the vaccination campaign in the hospital, but among this 16 were fully vaccinated and 35 were un-vaccinated. These were the study cohorts, i.e., infected vaccinated cohort and infected unvaccinated cohort. None of the healthcare professionals tested positive during the time gap between the first and second dose. On comparing the mean age between these two groups, the infected vaccinated cohort was slightly older (35.07 ± 10.38) than the infected unvaccinated group (31.97 ± 5.7). A female preponderance was observed in both -infected unvaccinated and infected vaccinated cohorts i.e., 97.14% and 75% respectively which was significant (p=0.014). A detailed description of the study cohorts with the significance level is depicted in Table 2.

**Table 2:** Demographic characteristics of the study cohorts.

| Table 2: Demographic characteristics of the study cohorts |  |  |  |
| --- | --- | --- | --- |
| Characteristics | Vaccinated & Infected (n=16) | Unvaccinated & Infected (n=35) | P value (0.05) |
| Mean Age (SD) | 35.07 (±10.38) | 31.97 (±5.7) |  |
| Gender |  |  |  |
| Female | 12 (75%) | 34 (97.14%) | p=0.014 |
| Male | 4 (25%) | 1 (2.85%) |  |
| Job Stratum |  |  |  |
| Doctors | 1 (6.25%) | 0 | p=0.029 |
| Nurses | 7 (43.75%) | 27 (77.14%) |  |
| Pharmacists | 2 (12.5%) | 0 |  |
| Lab & Diagnostics | 2 (12.5%) | 7 (20%) |  |
| Housekeeping | 1 (6.25%) | 1 (2.85%) |  |
| Physiotherapy | 1 (6.25%) | 0 |  |
| Medical Records Department | 1 (6.25%) | 0 |  |
| Maintenance | 1 (6.25%) | 0 |  |
| Exposure Grade |  |  |  |
| Direct Patient Contact | 11 (68.75%) | 34 (97.15%) | p= 0.029 |
| No Direct Patient Contact | 5 (31.25%) | 1 (2.85%) |  |

Healthcare team members with a greater likelihood for direct positive contact i.e., nurses were the most infected among both the cohorts. Infected vaccinated and infected unvaccinated HCWs differ significantly by age, sex, department, and their contact exposure grade. The median time between the second dose and the laboratory-confirmed COVID-19 infection was 65 (IQR: 20 -91 days). Among the 16 vaccinated healthcare workers who tested positive, all of them were symptomatic with symptoms ranging from fever, chills, sore throat, headache, and cough. None of these individuals required hospitalization.

The multivariate analyses using the Cox proportional hazard model revealed that the age of the healthcare worker (P = 0.856, HR = 0.994), sex (P = 0.435, HR = 1.290), and contact exposure status (P = 0.092, HR = 0.362) of the study cohorts, are not significant factors for getting infected after being vaccinated.

While the stepwise binary logistic regression analysis revealed a significant relationship exists between the incidence of infection and being unvaccinated (P = 0.001, OR = 4.278)., it also proves that age (P = 0.814, OR = 1.006) and sex (P = 0.407, OR = 0.605) of the individuals are not significant independent factors for getting infected.

Table 3 describes the percentage of infected in the vaccinated group as 6.58% and that in the unvaccinated group as 43.75%. On analyzing the vaccine effectiveness, it was calculated to be 84.95% (Cl =95; p<0.05). The Absolute Risk Reduction (ARR) was found to be 37.17% and the baseline risk to be 43.75% therefore, Relative Risk Reduction (RRR) was obtained using ARR/ Baseline risk that is 84.96%. This conveys that, completing the two-dose cycle of vaccination decreased the hazard of testing positive against the SARS-CoV-2 by 84.96 % when compared with the unvaccinated individuals.

**Table 3:** 2×2 contingency table to calculate the vaccine effectiveness.

| **Table 3: 2x2 contingency table to calculate the vaccine effectiveness** |  |  |
| --- | --- | --- |
|  | **COVID -19 Infected** | **COVID -19 Not Infected** |
| **Vaccinated** | 16 | 227 |
| **Not Vaccinated** | 35 | 46 |

The highest number of healthcare workers getting infected with COVID-19 in May 2021 depicts the peak incidence of the second wave. The breakthrough infections post-vaccinations occurred after February, 2021. The linear forecast (dotted line) graph (Figure 2), shows that the probability of getting infected in vaccinated HCPs is comparatively less than the unvaccinated group.

**Figure 2:**
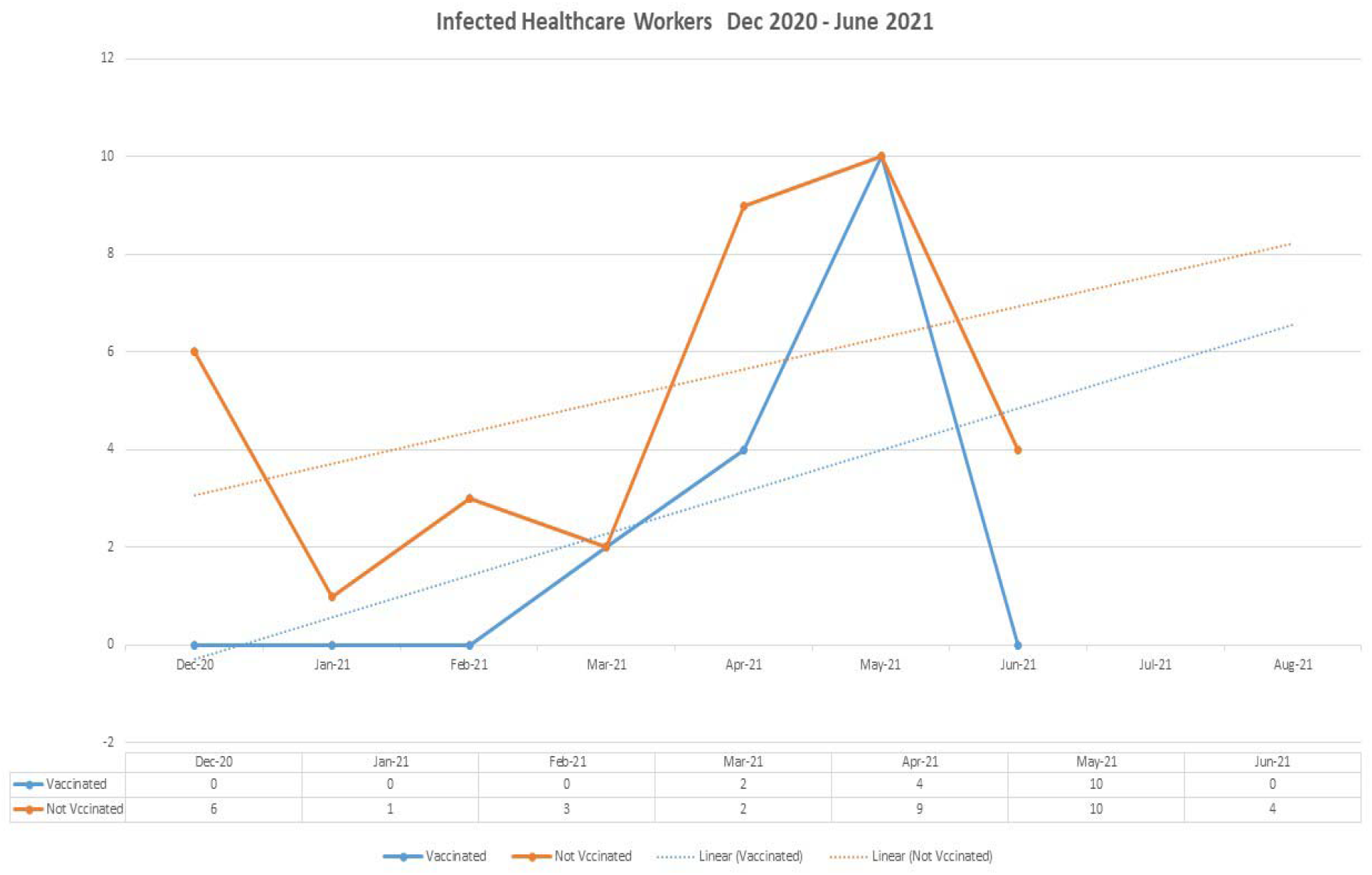
Incidence of breakthrough infections in the vaccinated and unvaccinated cohorts plotted against the respective month along with the linear forecast (dotted line).

## Discussion

ChAdOx1 nCoV-19 vaccine had proven their efficacy of 70·4% (95% CI 43·6–84·5) in post-hoc analysis on SARS-CoV-2, B.1.1.7 variant [7]. But, the real-time analysis of the breakthrough infections after completing the full course of the vaccine is crucial in determining the potency and effectiveness of the vaccine. Our study conducted in a secondary care hospital in the South Indian state of Kerala has pointed out the effectiveness of ChAdOx1 nCoV-19 vaccine among the healthcare workers who were continuously exposed to the COVID-19 positive patients and their close contacts.

Completing the full course of the vaccination decreased the risk of infection with the SARS-CoV-2 virus by 84.96% which is lower than the 91.8 – 94.9% in the VIN-WIN cohort [12] and the study done by Benal JL et.al i.e, 89% (85% to 93%) [13].

The breakthrough infection rate among the vaccinated health care workers in the previous studies was found to be 18.65% and 13.3% [14, 15] and it is 6.58 % in our study. Comparison with other studies is mentioned in Table 4.

**Table 4:** A comparison of study outcomes with the results from similar studies.

|  |  |  |  |  |
| --- | --- | --- | --- | --- |
| **Parameters** | **Our study** | **Subhadeep Ghosh et.al. [12]** | **Pragya Sharma et.al. [9]** | **Tyagi K et. al. [15]** |
| **Study site (Country)** | India | India | India | India |
| **Study population** | Healthcare workers | Indian Armed Forces | Healthcare workers | Healthcare workers |
| **Median/Mean days of Infection after the second dose of vaccine** | 65 days (IQR: 21 - 91) | 31 days (95% CI:30-33) | 47 days (Range: 28.5 - 55) | 34.8 days (Range: 2- 51) |
| **Incidence of breakthrough infection** | 6.58% | 26.2% | 11.3% | 16.8% |

The mean age of patients with breakthrough infections was 35.07 (±10.38) years and most of the vaccinated individuals, as well as the, vaccinated infected individuals were females i.e, 79.83% and 75% respectively similar to the findings of Sabnis R et.al [17].

In our study, the median days between the second dose and being tested positive for SARS-CoV-2 was 65 days which is much longer than reported in all the previous studies, which ranged from 5 days or 47 days [9, 12, 15, 16]. Also, none of the vaccinated HCWs who tested positive for SARS-CoV-2 were hospitalized, and there were no deaths, which in turn proves that vaccination reduces hospitalization by 100% which is more than the results observed in a study conducted in Scotland (88%) [18] and England (43%) [19]. Thus, we ascertain and uphold that the ChAdOx1 nCoV-19 vaccine is effective in protecting healthcare workers.

The symptomatic profile of the infected vaccinated HCWs was found similar to that study from Siloam Teaching Hospital, Indonesia [16]. Nurses (43.75%) and the staffs with more positive patient contacts (68.75%) were more infected among the vaccinated samples, which agree with the fact that more infection control practices have to be followed in the hospital settings [20].

Hesitancy against COVID-19 vaccine [33.75% (27)], being the major reason for being unvaccinated and the same trend was observed worldwide [21]. Other reasons include breastfeeding and pregnancy, which might be due to the reason that only very limited studies regarding the safety of vaccines in these categories were conducted during the initial phase of vaccination.

Limitations of the study is that this is a monocentric, observational study design with a small sample size and also the inability to adjust the confounding variables. Also, the study doesn’t compare the efficacy of various other vaccines available in India. Another limitation is that we don’t evaluate the immunogenicity of the vaccinated individuals and the results address the short-term effect of the vaccine. We need multi-national studies with a large sample size to provide more accurate results.

Breakthrough infections among the healthcare workers amid vaccination campaigns remain a concern for all authorities worldwide. As plotted in the dotted linear forecast (Figure 2), there is an increased risk of the third wave of COVID-19 infection in the coming months. So, the basic infection control measures including face masking, hand hygiene, social distancing, and quarantine protocols should be followed even if vaccinated, to prevent the risk of breakthrough infections.

## Data Availability

The data stated in the manuscript is available to all the authors.

## Abbreviations

ChAdOx1 nCoV-19: Chimpanzee (Ch) Adenovirus-vectored vaccine (Ad), developed by the University of Oxford (Ox)to stimulate an immune response to novel coronavirus (nCoV-19) first identified in 2019.
HCWs: Healthcare Workers
HCP: Healthcare Professional
HR: Hazard Ratio
IQR: Inter Quartile Range
OR: Odds Ratio

## Author Contribution

Design of research study (AI, JK), data acquisition (AI, JJ, LE), data analysis (AI), interpretation of results (AI, JJ), writing the manuscript (JJ).

